# Baseline and treatment-emergent bedaquiline resistance in drug-resistant tuberculosis: A systematic review and meta-analysis

**DOI:** 10.1101/2023.08.07.23293687

**Authors:** Rubeshan Perumal, Neda Bionghi, Camus Nimmo, Marothi Letsoalo, Matthew J. Cummings, Madeleine Hopson, Allison Wolf, Shamim Al Jubaer, Nesri Padayatchi, Kogieleum Naidoo, Michelle H. Larsen, Max O’Donnell

## Abstract

**Rationale:** Bedaquiline is a novel antimycobacterial agent for drug-resistant tuberculosis (DR-TB) and is classified as a World Health Organization (WHO) Group A drug due to its excellent clinical efficacy, high bactericidal activity, and potent sterilizing effect. Baseline and treatment-emergent bedaquiline resistance have been described but prevalence and incidence have not been reported, leading to gaps in the knowledge required to design strategies to optimize MDR-TB clinical outcomes and prevent the amplification of bedaquiline resistance.

**Methods:** We performed a systematic review and meta-analysis to estimate the frequency of, and mutations associated with, baseline and acquired (treatment-emergent) bedaquiline resistance in clinical *Mtb* isolates. Pooled estimates of bedaquiline resistance were generated by proportional meta-analysis in R version 4.2.2 using dmetar, metafor and meta packages. Resistance associated variants associated with prevalent and incident bedaquiline resistance were identified.

**Results:** Data from 14 studies were included; 14 and 9 studies reported on pre-treatment and acquired bedaquiline resistance, respectively. The pooled prevalence of pre-treatment bedaquiline resistance was 2.4% (95% CI 1.7 – 3.5), with significant heterogeneity across all studies (*I*^2^ 66%, p<0.01). The pooled prevalence of treatment-emergent bedaquiline resistance was 2.1% (95% CI 1.4 - 3.0), with no significant heterogeneity across the included studies (*I*^2^ 0%, p=0.97).

**Discussion:** We found a concerning frequency of bedaquiline resistance present at baseline and acquired during treatment. Urgent strategies are required to mitigate further resistance to this crucial drug.

---

Bedaquiline is a novel antimycobacterial agent for drug-resistant tuberculosis (DR-TB) and is classified as a World Health Organization (WHO) Group A drug due to its excellent clinical efficacy, high bactericidal activity, and potent sterilizing effect.(1) The introduction of bedaquiline into treatment regimens has enabled short-course all-oral multidrug-resistant TB (MDR-TB) regimens and the shortening of drug-susceptible TB treatment.(2, 3)

Bedaquiline targets F_1_F_0_-ATP synthase to impair *Mycobacterium tuberculosis* (*Mtb)* ATP synthesis and exerts other incompletely characterized bactericidal effects.(4) Variants in the target *atpE* and *atpB* genes and off-target mutations in *mmpR5, mmpL5*, and *pepQ* have been associated with bedaquiline resistance.(5, 6) We performed a systematic review and meta-analysis to estimate the frequency of, and mutations associated with, baseline and acquired (treatment-emergent) bedaquiline resistance in clinical *Mtb* isolates.

The study protocol was registered in PROSPERO (CRD42022346547) and the PRISMA guidelines were followed for reporting of the review methods and findings. Systematic searches of MEDLINE/PubMed, Cochrane Central Register of Clinical Trials, and EMBASE were conducted through February 2023 for publications on phenotypic resistance of bedaquiline. We included studies which reported clinical *Mtb* isolates with bedaquiline resistance via minimum inhibitory concentration (MIC) values from patients with at least rifampicin-resistant TB. Given the suboptimal positive predictive value of resistance-associated variants for phenotypic resistance, our study only evaluated phenotypic resistance as defined by MIC thresholds. We excluded studies with MIC cutoffs inconsistent with WHO cutoffs, *in vitro Mtb* isolates not obtained from patients, or ≤3 patients/isolates. Phenotypic bedaquiline resistance was defined by critical concentrations of 1⍰μg/ml by MGIT method or 0.25⍰μg/ml by broth microdilution or 7H11 agar proportion method. Acquired bedaquiline resistance was defined as the absence of phenotypic resistance before treatment and demonstration of phenotypic resistance on at least one occasion during bedaquiline treatment (Table S1). Publication bias was evaluated using funnel plots and methodological quality was assessed by the tool proposed by Hoy *et al*.(7) We performed a proportional meta-analysis in R version 4.2.2 using dmetar, metafor and meta packages. Paired MIC and resistance-associated variant (RAV) data were presented by scatterplots. Detailed methods are available in the Supplementary File.

The systematic search identified 180 articles for assessment: 37 were duplicates, 47 did not meet inclusion criteria, and 82 were excluded because they were in vitro studies, review papers, case reports, used non-standardized MIC thresholds, did not distinguish between baseline and treatment-emergent resistance, contained overlapping data with another study, or had no retrievable full text (**Figure S1**). The remaining 14 studies were included, comprising four randomized controlled trials and 10 cohort studies, emanating from five continents (**Table S2**). (5, 6, 8-19) In both baseline and during-treatment analyses there was evidence of significant positive publication bias. For the baseline analysis, 13 studies were rated as low risk of bias, and one study as moderate risk of bias. For the during-treatment analysis, three studies were rated as low risk of bias, and six studies as moderate risk of bias (**Tables S3 and S4**).

Fourteen studies, with a pooled sample of 9975 isolates, contributed to the estimate of baseline bedaquiline resistance. Nine studies, with a pooled sample of 1912 isolates, contributed to the estimate of treatment-emergent bedaquiline resistance. The pooled prevalence of baseline bedaquiline resistance was 2.4% (95% CI 1.7 – 3.5), with significant heterogeneity across all studies (*I*^2^ 66%, p<0.01) (**Figure 1, Panel A**). The pooled prevalence of treatment-emergent bedaquiline resistance was 2.1% (95% CI 1.4 - 3.0), with no significant heterogeneity across the included studies (*I*^2^ 0%, p=0.97) (**Figure 1, Panel B**). In the sensitivity analyses, no between-group differences were seen based on the quality of the studies (low versus moderate risk of bias).

**Figure 1.**
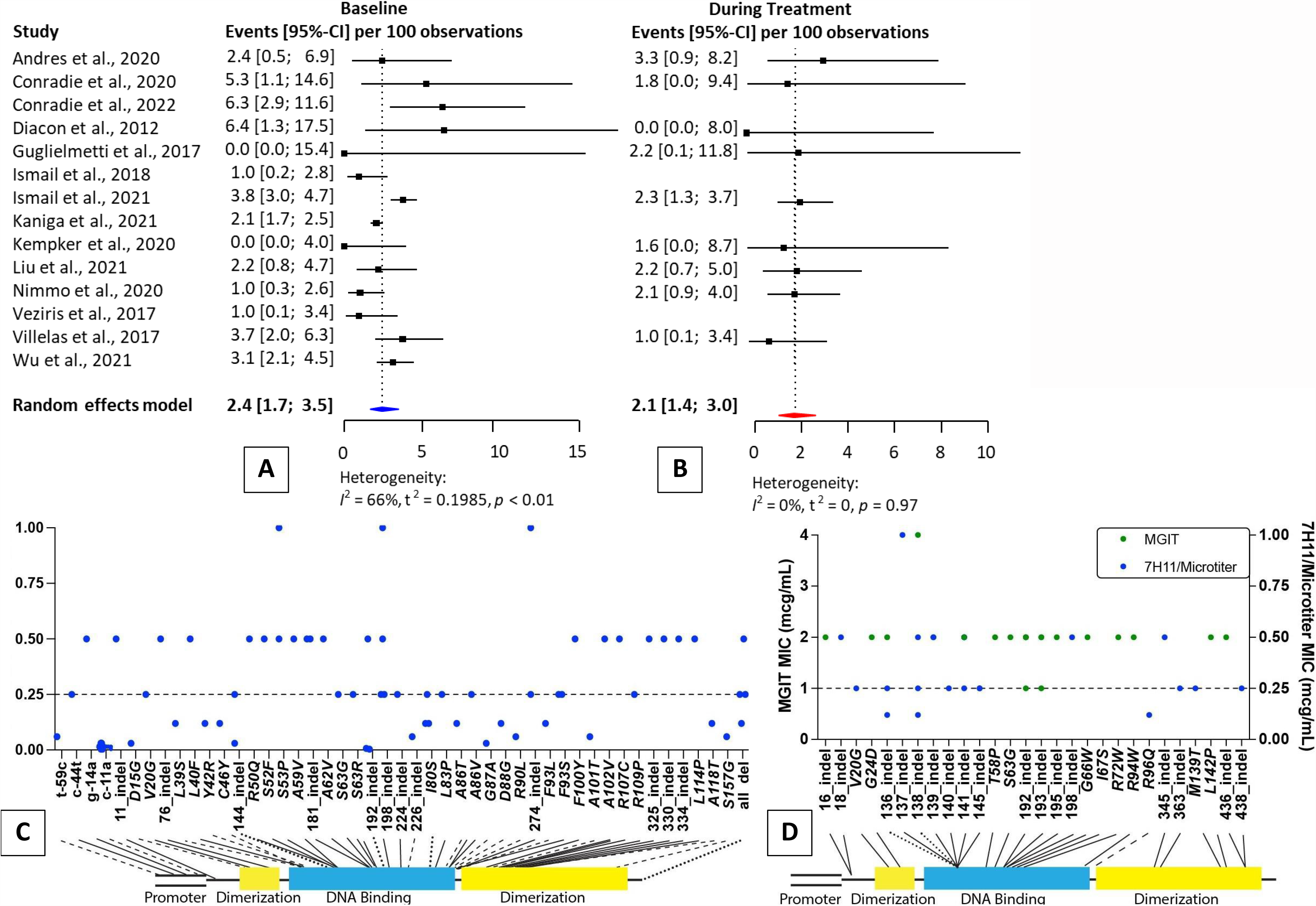
Pooled prevalence of baseline (**Panel A**) and treatment-emergent (**Panel B**) bedaquiline resistance in patients with drug-resistant TB. Baseline (**Panel C**) and treatment-emergent (**Panel D**) resistance-associated variants and associated minimum inhibitory concentrations. Solid black lines represent intermediate or resistant MICs, dashed lines represent susceptible MICs, and dotted lines are RAVs that are associated with both susceptible and intermediate/resistant MICs.

We identified 141 RAVs across all isolates in the included studies, comprising 71 unique RAVs at 58 unique sites. Four RAVs were mapped to the promoter region of the Mtb genome, 19 were mapped to the dimerization domains, and 42 were mapped to the DNA binding region. The complete list of RAVs and associated MICs is presented by scatterplot (**Figure 1, Panels C and D**). Treatment-emergent RAVs were significantly more likely to be associated with phenotypic resistance than pre-treatment RAVs [21/35 (60.0%, 95% CI 42.9 – 77.7) vs 24/97 (24.7%, 95% CI 15.8 – 33.7)].

The high degree of heterogeneity and publication bias suggests that our pooled estimate of baseline bedaquiline resistance (2.4%, 95% CI 1.7 – 3.5) may overestimate the true prevalence in clinical settings. However, the two included studies which involved comprehensive national surveillance of all patients initiating treatment for MDR-TB produced reliable estimates of pre-treatment bedaquiline resistance of 3.2% and 3.6% in Germany and South Africa, respectively.(8, 17) These levels of pre-treatment bedaquiline resistance likely reflect high rates of person-to-person community transmission of bedaquiline-resistant TB.

We found a concerning pooled prevalence of acquired bedaquiline resistance during treatment – 2.1% (95%CI 1.4 –3.0%). Our estimate is concordant with a previous systematic review which reported a frequency of phenotypic acquired bedaquiline resistance of 2.2%.(20) The low heterogeneity among included studies reflects the systematic evaluation of serial cultures from all patients treated for MDR-TB, ranging from weekly to alternate months until culture conversion. This approach avoided the spuriously high estimates of acquired bedaquiline resistance reported in studies that only performed serial phenotypic DST in patients with evidence of treatment failure.(21)

As *mmpR5 (Rv0678)* mutations are associated with cross-resistance to clofazimine and bedaquiline, prior exposure to clofazimine may have contributed to the high levels of pre-treatment bedaquiline resistance identified in settings with limited prior use of bedaquiline. As a WHO group B drug, clofazimine has been increasingly used as part of shortened MDR-TB regimens globally. Bedaquiline has a relatively long elimination half-life, estimated at around 5.5 months, rendering it particularly vulnerable to acquired resistance when treatment is interrupted. For this reason, high levels of adherence support, patient tracking, and robust novel treatment regimens with a high barrier to resistance are necessary to mitigate the risk of acquired resistance. In addition, bedaquiline exhibits sub-optimal penetration into caseous necrotic lesions exposing it to intralesional PK-PD mismatch.(22) Few MDR-TB drugs offer adequate protection against bedaquiline resistance, which is an increasingly important consideration when constructing an MDR-TB regimen.(8)

Bedaquiline received US Food and Drug Administration accelerated approval in 2012 and by 2014 – the first case of bedaquiline-resistant TB was reported in a Tibetan refugee following treatment with a bedaquiline-containing regimen.(23) Between 2015 and 2019, there were an estimated 300 cases of bedaquiline-resistant TB cases in South Africa alone.(17) We are reminded that the prevalence of rifampicin resistance was initially ∼12 per 1000 patients shortly after roll out of rifampicin, but amplified to ∼300 per 1000 patients currently.(24, 25) A Markov decision modelling exercise estimated the prevalence of bedaquiline resistance increasing to 588 per 1000 patients with more widespread bedaquiline rollout.(26)

As we prepare for the mass rollout of bedaquiline-containing short-course regimens for the treatment of MDR-TB, National TB Programmes will require substantial strengthening to avoid escalating levels of bedaquiline resistance, ensuing surges in treatment failure and relapse, a growing dependence on complex salvage regimens, and treatment destitution. Central to reducing the threat of bedaquiline-resistant TB epidemic is the need for a rapid diagnostic test to detect bedaquiline resistance. Research to better understand the genetics of resistance as well as investment in rapid phenotypic methods such as microscopic observation of drug susceptibility and reporter mycobacteriophages may be required. To date, the development of a rapid molecular diagnostic assay has been severely constrained by the poor phenotypic-genotypic concordance of resistance profiling for bedaquiline. In parallel, as bedaquiline-based regimens are implemented, national and supra-national bedaquiline resistance surveillance activities must be escalated as a critical early warning system.

## Data availability

Materials related to this review are available from the authors upon request.

