## Supplementary File for "Baseline and treatment-emergent bedaquiline resistance in drug-resistant tuberculosis: A systematic review and meta-analysis"

**Search strategy and study selection**

Systematic searches of MEDLINE/PubMed, Cochrane Central Register of Clinical Trials, and EMBASE were conducted from inception until October 2022 for publications that reported on phenotypic resistance of bedaquiline. The search strategy included terms in titles and abstracts: “Bedaquiline AND resistan*”, “TMC207 AND resistan*”, “[Bedaquiline AND (“RAV” OR “drug-resistant tuberculosis” OR “Rv0678” OR “atpE” OR “pepQ” OR “mmpl5” OR “mmpR” OR “Rv2535c” OR “Rv1305” OR “minimum inhibitory concentration”)]. Concepts were exploded to include all Medical Subject Headings (MeSH) subheadings. A manual search of all pertinent articles from the reference list of identified articles was conducted to identify additional articles not found by the electronic search. Publications were limited to human studies and the English language (or translated to English).

We included all studies which reported the number of isolates or patients with bedaquiline resistance via minimum inhibitory concentration (MIC) values in MTB isolates obtained from patients with resistance to at least rifampicin. Studies that were not in English (or translated to English), had MIC cutoffs not consistent with the WHO cutoffs (critical concentrations of 1 μg/ml by MGIT method or 0.25 μg/ml by broth microdilution or 7H11 agar proportion method), were exclusively *in vitro* (i.e., MTB isolates not obtained from patients), did not distinguish between baseline and treatment-emergent resistance, or were case reports/case series with less than or equal to 3 patients/isolates were excluded.(1)

Baseline bedaquiline resistance was defined as the presence of phenotypic bedaquiline resistance in a sputum sample obtained prior to the initiation of bedaquiline-containing treatment. Acquired resistance was defined as the presence of phenotypic bedaquiline resistance in a sputum sample obtained during treatment with a bedaquiline-containing regimen, in the absence of phenotypic bedaquiline resistance in a baseline/pre-treatment sample. (**Table S1**)

**Table S1**. Calculation of the frequency of baseline and acquired bedaquiline resistance.

| **Measure** | **Numerator** | **Denominator** |
| --- | --- | --- |
| **Baseline Phenotypic Resistance** | Number of patients with **MIC‑based resistance** in a baseline/pre-treatment isolate | Number of patients **with rifampicin-resistant tuberculosis** |
| **Acquired Phenotypic Resistance** | Number of patients with **MIC‑based resistance** on an isolate obtained during treatment with a bedaquiline-containing regimen | Number of patients with rifampicin-resistant tuberculosis **treated with bedaquiline** AND **without baseline MIC-based bedaquiline resistance** |

**Data extraction, quality assessment, and statistical analysis**

Two reviewers (NB and MH) independently screened all identified articles for inclusion, and any disagreement was settled through discussion and consensus. Duplicate studies of the same cohort/data were excluded. The following data elements were extracted onto a standardized data extraction form: author, year of publication, study setting, sample size, number of isolates, bedaquiline MICs (per isolate, where available) and treatment data (where available). The authors of the relevant studies were contacted and requested to provide further information if the summary statistics provided in the published manuscript were not adequate for our analysis.

For each included study, we determined the proportion of patients with baseline bedaquiline resistance and acquired bedaquiline resistance. The following definitions were used: MDR-TB was defined as TB resistant to isoniazid and rifampicin; *RR-TB* was defined as TB resistant to at least rifampicin; *TB treatment* was defined as treatment with a regimen that included bedaquiline; *phenotypic bedaquiline resistance* was defined by critical concentrations of 1 μg/ml by MGIT method and 0.25 μg/ml by broth microdilution or 7H11 agar proportion (AP) method; *acquired bedaquiline resistance* was defined as the absence of phenotypic bedaquiline resistance before treatment and the demonstration of phenotypic resistance on at least one occasion during treatment. The methodological quality of the included studies was assessed using the Risk of Bias Assessment proposed by Hoy et al. Publication bias was evaluated by visual inspection of funnel plots, visual inspection of Doi plots, and LFK indices. An LFK index between -1 and +1 was consistent with the absence of significant asymmetry, while an LFK <-1 or >+1 was reflective of significant asymmetry.

We performed a proportional meta-analysis (PMA) on data for DR-TB patients with bedaquiline resistance at baseline (i.e. pre-treatment resistance) and patients who acquired bedaquiline resistance on treatment (i.e. treatment-emergent resistance). The conduct of the two PMAs fitted a generalized linear mixed model to account for the differences of the effect sizes of the included studies. Data analysis for this study were carried out in R version 4.2.2 using dmetar, metafor and meta packages following the work of Barker et al. (2021) and Harrer et al. (2021).(2, 3) Based on the guidelines by Barker et al. (2021) on the conduct of PMA, our analyses present study-specific proportion estimates with their respective 95% confidence intervals (95% CIs) together with the pooled estimate (proportion and 95% CI). Prevalence and their 95% CI are presented as per 100 observations. The heterogeneity of estimates between studies was assessed with the *I^2^* statistic from Cochran Q. An *I^2^* > 30% and p<0.1 was considered reflective of significant heterogeneity. Paired MIC and RAV data, from studies which reported such data, were presented by scatterplots.

Records identified from:

PubMed (n = 114)

EMBASE (n = 42)

Cochrane (n = 24)

Records removed before screening:

Duplicates removed by Endnote (n = 37)

Titles and abstracts screening (n = 143)

Studies not included due to inclusion criteria: (n = 47)

Full-text articles assessed for eligibility (n = 96)

Studies excluded due to exclusion criteria (n = 82):

Irrelevant (n = 27)

Full text unavailable (n = 1)

Did not differentiate baseline and treatment-emergent resistance (7)

MIC not standardised (n = 7)

Case report/series (n = 8)

Review papers (n = 14)

In vitro studies (n = 15)

Duplicate data (n = 3)

Studies included in review and meta-analysis (n = 14)

**Figure S1.** PRISMA flow chart of literature search

**Table S2.** Characteristics of included studies(4-17)

| **Author, year** | **Setting** | **Period** | **Design** | **Sample Size** | **Total resistant** | **Baseline** | **Acquired** |
| --- | --- | --- | --- | --- | --- | --- | --- |
| Andres et al., 2020 | Europe | 2018-2019 | Cohort | 124 | 7 | 3 | 4 |
| Conradie et al., 2020 | South Africa | 2015-2017 | RCT | 57 | 4 | 3 | 1 |
| Conradie et al., 2022 | South Africa | 2017-2019 | RCT | 181 | 9 | 9 | -- |
| Diacon et al., 2012 | South Africa | 2007-2008 | RCT | 47 | 3 | 3 | 0 |
| Guglielmetti et al., 2017 | France | 2011-2013 | Cohort | 45 | 1 | 0 | 1 |
| Ismail et al., 2018 | South Africa | 2012-2014 | Cohort | 310 | 3 | 3 | -- |
| Ismail et al., 2021 | South Africa | 2015-2019 | Cohort | 2004 | 92 | 76 | 16 |
| Kaniga et al., 2021 | Multi-country | 2015-2019 | Cohort | 5036 | 104 | 104 | -- |
| Kempker et al., 2020 | Georgia | 2015-2017 | Cohort | 95 | 1 | 0 | 1 |
| Liu et al., 2021 | China | Not reported | Cohort | 277 | 11 | 6 | 5 |
| Nimmo et al., 2020 | South Africa | 2013-2019 | Cohort | 391 | 12 | 4 | 8 |
| Veziris et al., 2017 | France | 2014-2015 | Cohort | 209 | 4 | 2 | 2 |
| Villellas et al., 2017 | Belgium | 2009-2013 | RCT | 347 | 13 | 13 | -- |
| Wu et al., 2021 | China | 2008-2019 | Cohort | 898 | 28 | 28 | -- |

**Table S3.** Risk of Bias Assessment, baseline bedaquiline resistance.

| **Study** | **1** | **2** | **3** | **4** | **5** | **6** | **7** | **8** | **9** | **10 Summary** |
| --- | --- | --- | --- | --- | --- | --- | --- | --- | --- | --- |
| Andres et al., 2020 | 0 | 0 | 0 | 1 | 0 | 0 | 0 | 0 | 1 | 2 (Low risk) |
| Conradie et al., 2020 | 1 | 1 | 0 | 1 | 0 | 0 | 0 | 0 | 0 | 3 (Low risk) |
| Conradie et al., 2022 | 1 | 0 | 0 | 0 | 0 | 0 | 0 | 0 | 0 | 1 (Low risk) |
| Diacon et al., 2012 | 1 | 0 | 0 | 0 | 0 | 0 | 0 | 0 | 0 | 1 (Low risk) |
| Guglielmetti et al., 2017 | 1 | 0 | 0 | 1 | 0 | 0 | 0 | 0 | 1 | 3 (Low risk) |
| Ismail et al., 2018 | 0 | 0 | 1 | 1 | 0 | 0 | 0 | 0 | 1 | 3 (Low risk) |
| Ismail et al., 2021 | 0 | 0 | 0 | 1 | 0 | 0 | 0 | 0 | 0 | 1 (Low risk) |
| Kaniga et al., 2021 | 1 | 0 | 0 | 1 | 0 | 0 | 0 | 0 | 0 | 2 (Low risk) |
| Kempker et al., 2020 | 0 | 0 | 0 | 1 | 0 | 0 | 0 | 0 | 0 | 1 (Low risk) |
| Liu et al., 2021 | 1 | 0 | 0 | 0 | 0 | 0 | 0 | 0 | 0 | 1 (Low risk) |
| Nimmo et al., 2020 | 1 | 1 | 1 | 1 | 0 | 0 | 0 | 0 | 1 | 5 (Moderate risk) |
| Veziris et al., 2017 | 0 | 0 | 0 | 1 | 0 | 0 | 0 | 0 | 1 | 2 (Low risk) |
| Villellas et al., 2017 | 1 | 0 | 0 | 1 | 0 | 0 | 0 | 0 | 0 | 2 (Low risk) |
| Wu et al., 2021 | 0 | 0 | 0 | 1 | 0 | 0 | 0 | 0 | 0 | 1 (Low risk) |

1. Was the study’s target population a close representation of the national DR-TB population? (Yes =0, No=1)
2. Was the sampling frame a true or close representation of the target population? (Yes =0, No=1)
3. Was some form of random selection used to select the sample, OR, was a census undertaken? (Yes =0, No=1)
4. Was the likelihood of non-response bias minimal? (Yes =0, No=1)
5. Were data collected directly from the subjects (as opposed to a medical records)? (Yes =0, No=1)
6. Were acceptable case definitions used in the study? (Yes =0, No=1)
7. Were reliable and accepted diagnostic methods for diagnosing bedaquiline resistance utilised? (Yes =0, No=1)
8. Was the same mode of data collection used for all subjects? (Yes =0, No=1)
9. Were the numerator(s) and denominator(s) for the calculation of the proportion of bedaquiline resistance appropriate? (Yes =0, No=1)
10. Summary on the overall risk of study bias (Low risk = 0-3, Moderate risk = 4-6, High risk = 7-9)

**Adapted from:** Hoy D, Brooks P, Woolf A, Blyth F, March L, Bain C, et al. Assessing risk of bias in prevalence studies: modification of an existing tool and evidence of interrater agreement. J Clin Epidemiol. 2012;65: 934-939.(18)

**Table S4.** Risk of Bias Assessment, acquired bedaquiline resistance.

| **Study** | **1** | **2** | **3** | **4** | **5** | **6** | **7** | **8** | **9** | **10 Summary** |
| --- | --- | --- | --- | --- | --- | --- | --- | --- | --- | --- |
| Andres et al., 2020 | 1 | 1 | 1 | 1 | 0 | 0 | 0 | 0 | 1 | 5 (Moderate risk) |
| Conradie et al., 2020 | 1 | 1 | 0 | 1 | 0 | 0 | 0 | 0 | 1 | 4 (Moderate risk) |
| Diacon et al., 2012 | 1 | 1 | 0 | 1 | 0 | 0 | 0 | 0 | 1 | 4 (Moderate risk) |
| Guglielmetti et al., 2017 | 1 | 1 | 1 | 1 | 0 | 0 | 0 | 0 | 1 | 5 (Moderate risk) |
| Ismail et al., 2021 | 0 | 0 | 0 | 1 | 0 | 0 | 0 | 0 | 0 | 1 (Low risk) |
| Kempker et al., 2020 | 0 | 0 | 0 | 1 | 0 | 1 | 0 | 0 | 1 | 3 (Low risk) |
| Liu et al., 2021 | 0 | 0 | 0 | 1 | 0 | 0 | 0 | 0 | 0 | 1 (Low risk) |
| Nimmo et al., 2020 | 1 | 1 | 1 | 1 | 0 | 0 | 0 | 0 | 1 | 5 (Moderate risk) |
| Veziris et al., 2017 | 0 | 1 | 1 | 1 | 0 | 0 | 0 | 0 | 1 | 4 (Moderate risk) |

1. Was the study’s target population a close representation of the national DR-TB population? (Yes =0, No=1)
2. Was the sampling frame a true or close representation of the target population? (Yes =0, No=1)
3. Was some form of random selection used to select the sample, OR, was a census undertaken? (Yes =0, No=1)
4. Was the likelihood of non-response bias minimal? (Yes =0, No=1)
5. Were data collected directly from the subjects (as opposed to a medical records)? (Yes =0, No=1)
6. Were acceptable case definitions used in the study? (Yes =0, No=1)
7. Were reliable and accepted diagnostic methods for diagnosing bedaquiline resistance utilised? (Yes =0, No=1)
8. Was the same mode of data collection used for all subjects? (Yes =0, No=1)
9. Were the numerator(s) and denominator(s) for the calculation of the frequency of bedaquiline resistance appropriate? (Yes =0, No=1)
10. Summary on the overall risk of study bias (Low risk = 0-3, Moderate risk = 4-6, High risk = 7-9)

**Adapted from:** Hoy D, Brooks P, Woolf A, Blyth F, March L, Bain C, et al. Assessing risk of bias in prevalence studies: modification of an existing tool and evidence of interrater agreement. J Clin Epidemiol. 2012;65: 934-939.(18)
